# Diminished capacities to make treatment decision for COVID-19 vaccination in schizophrenia

**DOI:** 10.1101/2022.01.11.22269070

**Authors:** Stéphane Raffard, Sophie Bayard, Margot Eisenblaetter, Philippe Tattard, Jérôme Attal, Yasmine Laraki, Delphine Capdevielle

## Abstract

Recent evidence suggests that schizophrenia patients are at high risk for severe COVID-19 and should be prioritized for vaccination. However, impaired decision-making capacities could negatively affect the uptake of COVID-19 vaccination in this population. Competence to consent to COVID-19 vaccination was assessed in 80 outpatients with schizophrenia. Using the MacArthur Competence Assessment Tool for Treatment, 56.3% of the sample were classified as having diminished mental capacity. Poor performance was associated with lower vaccination rates, poorer cognition and higher level of psychotic symptoms. Developing interventions for enhancing informed consent for vaccination is urgent within this population.

## Introduction

The worldwide pandemic caused by the Covid-19 (SARS-CoV-2) has strongly impacted people’s lives with dramatic economical and global health consequences. To face this unprecedented health crisis, different strategies including population confinement have been implemented worldwide. Among the strategies, the development of COVID-19 vaccination was an important and fundamental tool to limit the propagation of the pandemic; particularly in high-risk populations such as the elderly, individuals with cardiovascular or respiratory diseases, or people with mental health illnesses.

Several nationwide cohort studies and analyses using electronic health records revealed that having a mental health disorder was associated with increased and more severe COVID-19 related outcomes including mortality [1]. This was especially the case for severe mental health illnesses such as schizophrenia spectrum disorders. Consequently, individuals with schizophrenia should be prioritised in vaccine allocation strategies [2]. However, several important barriers could negatively affect the uptake of a COVID-19 vaccine in this population, including impaired decision-making capacitity. Indeed, impaired decision-making ability has been repeatedly observed in schizophrenia patients. Using both experimental tasks, such as the Iowa Gambling Task and clinical tools such as the MacArthur Competence Assessment Tool for Clinical Research (MacCAT-CR) or for Treatment (MacCAT-T), studies have repeatedly found altered decision-making capacities in schizophrenia patients, mainly due to cognitive deficits and psychotic symptoms [3,4]. Ability to make treatment decision has been proposed as one of the most impaired decision-making capacity in schizophrenia [5]. Using instruments, like the MacCAT-T, which is considered the standard measure for assessment of competence, it was found that 10% to 52% of people with schizophrenia were classified as being impaired in decisional capacity for treatment [6].

Nevertheless, to our knowledge, no studies have explored competence to consent to vaccination, particularly COVID-19 vaccination in people with schizophrenia. The aims of this investigation were to examine the competence to consent to COVID 19 vaccines in individuals with schizophrenia and 2) the clinical determinants of competence to consent to COVID 19 vaccines in patients. We hypothesized, first, that a high proportion of patients have reduced competency and second, that competence to consent will be inversely related with the severity of psychotic symptoms and positively, to cognitive functioning.

## Methods

### Participants

We assessed 80 outpatients with schizophrenia in the University Department of Adult Psychiatry of Montpellier, France between April 2021 to October 2021. Inclusion criteria were: (a) age between 18 and 60 years, (b) a DSM-5 diagnosis of schizophrenia, (c) adequate proficiency in French. Exclusion criteria for all participants were: (a) known neurological disease, (b) history of learning disability/developmental disorder.

### Measures

**Competence to consent to COVID-19 vaccination** was assessed with the French version of the MacCAT-T [7,8] The patient’s understanding of the COVID-19 disorder and treatment-related information (Understanding) was rated from 0 to 6; appreciation of the significance of that information for the patient, in particular the benefits and risks of treatment (Appreciation) was rated from 0 to 4; the reasoning ability of the patient (Reasoning) was rated from 0 to 8; and ability of the patient to express a choice between a vaccine against COVID-19 and an alternative treatment (Expressing a choice) was rated from 0 to 2. In order to reduce demands on the patient’s cognitive abilities, we used the MacCAT-T in the same way as Owen et al. [10] and compared vaccine for COVID-19 to no medical treatment, i.e., we did not offer an alternative treatment. Patients were divided into two groups based on ratings on the four subscales of the MacCAT-T and on the methodology of Elzakkers et al. [9] For every subscale a patient could rate poor (50% or less of the maximum rating on that subscale), intermediate (51–75% of the maximum rating) or good (76–100%). If a patient had a poor or intermediate rating on one or more of the four subscales, this patient was considered as having diminished mental capacity

### Clinical and cognitive assessment

Symptom severity was measured using the Positive and the Negative Syndrome Scale PANSS. For this study, four of the analytically derived PANSS factor component scores were taken into account: Total, General Psychopathology, Positive and Negative [10].

Global cognitive functioning was assessed with the Montreal Cognitive Assessment (MoCA) [11]. The MoCA is a brief measure of global cognitive function originally developed to detect mild cognitive impairment. Total score was the dependent variable.

### Procedure

Written informed consent was obtained from all participants prior to their inclusion in the study. Patients completed all measures in one session. All participants gave their written consent to participate. To reduce demand on the patient’s cognitive abilities, we used the MacCAT-T in the same way as Owen et al. and compared COVID-19 vaccine to no medical treatment, i.e., we did not offer an alternative treatment [6].

### Statistical analyses

Group comparison were made with the χ2 test for nominal variables and independent t-test for continuous variables. P values were 2-tailed, and significance was set at a P value less than .05. Analyses were performed using Jamovi software version 1.6.

## Results

The collected sociodemographic and clinical data are reported in Table 1. As documented in Table 1, 58.8% of patients had a full vaccine status. Diminished mental capacity was observed and ranged from 8.8% for the Expressing a choice dimension to 56.3% for the Understanding dimension.

**Table 1.** Sociodemographic and clinical characteristics of the sample

| Patients Characteristics | Value: mean (s.d.) [range] or % |  |  |  |  |
| --- | --- | --- | --- | --- | --- |
|  | All sample | Vaccinated | Unvaccinated | Statistics | P-value |
|  | *n* = 80 | *n* = 47 | *n* = 33 |  |  |
| Demographic variable |  |  |  |  |  |
| Age | 41.2 (13.2) | 42.7 (12.63) | 39.1 (13.97) | *t* = -1.20 | 0.23 |
| Gender, female | 30% | 36.2% | 21.2% | $\chi^2 = 2.07$ | 0.15 |
| Years of scholarship, mean | 12.6 (3.16) | 12.9 (3.3) | 12 (2.9) | *t* = -1.23 | 0.22 |
| Full vaccine status | 58.8% |  |  |  |  |
| Clinical variables |  |  |  |  |  |
| Patients under treatment | 100% |  |  |  |  |
| Duration of the disease | 16.8 (12.4) | 15.8 (12.5) | 18.2 (12.2) | *t* = 0.77 | 0.44 |
| PANSS<sup>a</sup> total | 62.1 (18.3) | 60 (18.7) | 66.2 (17.2) | *t* = 1.15 | 0.25 |
| PANSS positive | 13.2 (5.7) | 12.9 (6.39) | 13.9 (4.4) | *t* = 0.62 | 0.53 |
| PANSS negative | 16.3 (5.5) | 15.6 (5.52) | 17.6 (5.52) | *t* = 1.23 | 0.22 |
| PANSS general psychopathology | 32.6 (9.17) | 31.5 (8.96) | 34.6 (9.48) | *t* = 1.12 | 0.26 |
| Montral Cognitive Assessment | 23.5 (4.3) | 23.8 (4) | 23.2 (4.7) | *t* = -0.57 | 0.56 |
| MacCAT-T<sup>b</sup> |  |  |  |  |  |
| Understanding disorder | 56.3%<sup>c</sup> | 53.2%<sup>d</sup> | 60.6%<sup>d</sup> | $\chi^2 = 0.43$ | 0.51 |
| Appreciation | 32.5%<sup>c</sup> | 19.1%<sup>d</sup> | 51.5%<sup>d</sup> | $\chi^2 = 9.26$ | 0.002* |
| Reasoning | 31.3%<sup>c</sup> | 19.1%<sup>d</sup> | 48.5%<sup>d</sup> | $\chi^2 = 7.77$ | 0.005* |
| Expressing a choice | 8.8%<sup>c</sup> | 4.3%<sup>d</sup> | 15.2%<sup>d</sup> | $\chi^2 = 2.88$ | 0.09 |
| Comorbidities |  |  |  |  |  |
| Diabetes | 11.2% | 12.8%<sup>d</sup> | 9.1%<sup>d</sup> | $\chi^2 = 0.26$ | 0.60 |
| Pulmonary disease | 15% | 17%<sup>d</sup> | 12.1%<sup>d</sup> | $\chi^2 = 0.36$ | 0.54 |
| Cardiovascular disease | 6.2% | 8.5%<sup>d</sup> | 3%<sup>d</sup> | $\chi^2 = 0.99$ | 0.31 |

a. Positive and Negative Syndrome Scale
b. MacArthur Competence Assessment Tool for Treatment
c. Diminished mental capacity
d. Proportion among patients with diminished mental capacity on MacArthur Competence Assessment Tool for Treatment

Group comparisons indicated that diminished competence to consent to COVID-19 vaccination was related to a poorer global cognitive functioning for the Understanding (21.9 ± 4.6 *versus* 25.6 ± 2.7; Welch’s *t* = -4.35; *P* < 0.001; Cohen’s *d* = -0.95) and Appreciation (22 ± 5.1 *versus* 24.2 ± 3.7; Welch’s *t* = 1.95; *P* = 0.05; Cohen’s *d* = 0.77) MacCAT dimensions.

Higher level of psychotic symptoms, based on the PANSS scores, was related to diminished competence to consent, specifically for the Understanding dimension (55.4 ± 10.2 *versus* 67 ± 21.7; Welch’s *t* = 2.72; *P* = 0.01; Cohen’s *d* = 0.48) and the Appreciation dimension (58.8 ± 19.3 *versus* 69.8 ± 13.1; Welch’s *t* = -2.32; *P* = 0.02; Cohen’s *d* = -0.66).

Diminished competence to consent to COVID-19 vaccination on Appreciation and Reasoning of the MacCAT dimensions was associated with lower vaccination rates, respectively, *P* = 0.002 and *P* = 0.005.

No relationships were found between vaccination status, demographical, clinical, global cognitive functioning, and comorbidity variables (Table 1).

## Discussion

To our knowledge, this is the first study to investigate the question of competence to consent to COVID-19 vaccine and more broadly vaccination in individuals with schizophrenia. Our results revealed that in our clinical sample there was a high rate of reduced capacity to make decisions for COVID-19 vaccination. Most importantly the subgroup of patients with diminished capacity to consent had a lower vaccination rate than the group with persevered capacity, suggesting that diminished competence to consent might constitute a barrier for COVID-19 vaccination. Moreover, impaired competence to consent was associated with higher levels of psychotic symptoms and reduced cognitive abilities which is in accordance with previous studies about decision-making capacity for other treatments such as antipsychotics [4].

Despite accumulating evidence that people with schizophrenia are at high risk for severe forms of COVID-19 and long-term sequelae (including hospitalizations and mortality) a recent longitudinal study indicated that this population remains under-vaccinated against COVID-19 compared to the general population Several factors including costs, accessibility problems, and the absence of medical recommendations have been associated with lower COVID-19 vaccination rates in individuals with schizophrenia [12]. This study indicates that reduced competence to consent to vaccination may constitute a significant barrier to an efficient vaccination program in this population.

If there is preliminary evidence of the efficiency of targeted vaccination programs for people with mental disorders, our findings are of major importance in cases of vaccine refusal [13]. From an ethical and recovery point of view, autonomy and the right to self-determination are of fundamental importance for people with severe mental disorders. Clinicians faced with a patient’s refusal to be vaccinated should thus make an appropriate assessment of the individuals’ decision-making capacity by using tools such as the MacCAT-T. Importantly both uncertainty and unwillingness to be vaccinated against COVID-19 can be partly reduced by appropriate interventions integrating feedback, and providing clear information on the virus and disease itself [14]. In addition, interventions aiming at enhancing mental capacity for vaccination, which has been shown to be effective and feasible in this population, should be considered before implementing compulsory medical treatments [15, 16]. It is important that healthcare professionals understand the risks and benefits of covid-19 vaccination for people with schizophrenia, so that they can support them in reaching an informed decision. Indeed «In the end, it should be left to the individual to weigh the benefits and the risks, and to give informed consent for vaccination", even in individuals with severe mental disorders [2].

### Limitations

This study has limitations. The relative small sample size. This is a monocentric study in a sample of French individuals with schizophrenia.

## Data Availability

All data produced in the present study are available upon reasonable request to the authors

## Statement of Ethics

Participants in the study were treated in accordance with international ethical standards, including APA standards of ethics, and was approved by the hospital!s institutional review board (IRB ID : 202100768).

## Authors contributions

S.R. and D.C. designed the study. M.E., D.C. Y.L., J.A. and P.T. collected the data. S.B. conducted statistical analyses. S.R. wrote the first draft of the paper. All the authors were involved in discussing the findings and writing the manuscript. They all approved its final version.

## Data availability

The anonymised dataset is available from the corresponding author on reasonable request Declarations

## References

1. Fond G, Nemani K, Etchecopar-Etchart D, Loundou A, Goff DC, Lee SW, Lancon C, Auquier P, Baumstarck K, Llorca PM, Yon DK, Boyer L (2021). Association Between Mental Health Disorders and Mortality Among Patients With COVID-19 in 7 Countries: A Systematic Review and Meta-analysis. JAMA Psychiatry 78(11):1208–1217.

2. Mazereel V, Van Assche K, Detraux J, De Hert M (2021). COVID-19 vaccination for people with severe mental illness: why, what, and how? Lancet Psychiatry 8(10):860–861.

3. Betz LT, Brambilla P, Ilankovic A, Premkumar P, Kim MS, Raffard S, Bayard S, Hori H, Lee KU, Lee SJ, Koutsouleris N, Kambeitz J (2019). Deciphering reward-based decision-making in schizophrenia: A meta-analysis and behavioral modeling of the Iowa Gambling Task. Schizophr Res 204:7–15.

4. Spencer BWJ, Shields G, Gergel T, Hotopf M, Owen GS (2017). Diversity or disarray? A systematic review of decision-making capacity for treatment and research in schizophrenia and other non-affective psychoses. Psychol Med 47:1906–1922.

5. Appelbaum PS, Grisso T (1988). Assessing patients’ capacities to consent to treatment. N Engl J Med 319(25):1635–1638.

6. Owen GS, Szmukler G, Richardson G, David AS, Raymont V, Freyenhagen F, Martin W, Hotopf M (2013). Decision-making capacity for treatment in psychiatric and medical in-patients: cross-sectional, comparative study. Br J Psychiatry 2013203(6):461–467.

7. Grisso T, Appelbaum PS, Hill-Fotouhi C (1997). The MacCAT–T: a clinical tool to assess patients’ capacities to make treatment decisions. Psychiatr. Serv 48(11):1415–1419.

8. Raffard S, Lebrun C, Laraki Y, Capdevielle D (2021). Validation of the French Version of the MacArthur Competence Assessment Tool for Treatment (MacCAT-T) in a French Sample of Individuals with Schizophrenia: Can J Psychiatry 66(4):395–405

9. Elzakkers IFFM, Danner UN, Grisso T, Hoek HW, van Elburg AA (2018). Assessment of mental capacity to consent to treatment in anorexia nervosa: A comparison of clinical judgment and MacCAT-T and consequences for clinical practice. Int J Law Psychiatry 58:27–35

10. Kay SR, Fiszbein A, Opler LA (1987). The positive and negative syndrome scale (PANSS) for schizophrenia. Schizophr Bull 13(2):261–276.

11. Nasreddine ZS, Phillips NA, Bédirian V, Charbonneau S, Whitehead V, Collin I, Cummings JL, Chertkow H (2005). The Montreal Cognitive Assessment, MoCA: a brief screening tool for mild cognitive impairment. J Am Geriatr Soc 53(4):695–699.

12. Tzur Bitan D, Kridin K, Cohen AD, Weinstein O (2021). COVID-19 hospitalisation, mortality, vaccination, and postvaccination trends among people with schizophrenia in Israel: a longitudinal cohort study. Lancet Psychiatry. 8(10):901–908. doi: 10.1016/S2215-0366(21)00256-X.

13. Mazereel V, Vanbrabant T, Desplenter F, De Hert M (2021). COVID-19 vaccine uptake in patients with psychiatric disorders admitted to or residing in a university psychiatric hospital. Lancet Psychiatry 8:860–861.

14. Paul E, Steptoe A, Fancourt D (2021). Attitudes towards vaccines and intention to vaccinate against COVID-19: Implications for public health communications. Lancet Reg Health Eur 1:100012.

15. Dunn LB, Lindamer LA, Palmer BW, Schneiderman LJ, Jeste DV (2001). Enhancing comprehension of consent for research in older patients with psychosis: a randomised study of a novel consent procedure. Am J Psychiatry 158: 1911–1913.

16. Jeste DV, Palmer BW, Golshan S, Eyler LT, Dunn LB, Meeks T, Glorioso D, Fellows I, Kraemer H, Appelbaum PS (2009). Multimedia consent for research in people with schizophrenia and normal subjects: a randomized controlled trial. Schizophr Bull 35(4):719–729

